# Does Telemedicine Reduce health disparities? Longitudinal Evidence during the COVID-19 Pandemic in the US

**DOI:** 10.1101/2021.03.01.21252330

**Authors:** Ali Roghani, Samin Panahi

**Author notes:** Corresponding author: Ali Roghani.

## Abstract

**Objective:** The COVID-19 pandemic could be a significant health issue for the elderly population and those with pre-excising chronic condition. In response to the pandemic health care services have increased the use of telehealth medicine. The propose of this study is to examine factors associated with access to telemedicine before and after COVID-19 based on sociodemographic factors and type of chronic disease.

**Method:** We have used data from the Research and Development Survey (RANDS) at two different time points Data collection for the first wave occurred between June 9, 2020 and July 6, 2020 (n= 6786), second wave was between August 3, 2020 and August 20, 2020 (n=5972). Three questions have been asked from the participant: 1) did the provider offer telemedicine before the pandemic? 2) does the provider offer telemedicine during the pandemic? And 3) have the participants schedule telemedicine appointments?

**Result:** In both waves, 62 % of the participants reported providers did not have telemedicine services prior to the COVID-19 pandemic. However, we found a 22% increase in offering telemedicine in six first month of the COVID-19 pandemic. The finding shows almost no change in providing telemedicine between June and August. The data indicates just a 0.5% and 0.1% increase in accessing telemedicine, and scheduling in August than June, respectively. Patients older than 65 had higher access to telemedicine and had higher scheduling frequencies than other age groups, while they had the lowest access prior to the COVID-19. Blacks had the highest access to telemedicine services than other races (40%). Additionally, females, higher education, and living in metropolitan areas were associated with higher access and scheduling during the pandemic. There was a variation of access and scheduling in different chronic diseases, however, providers offered more remote services for those who diagnosed by diabetes.

**Conclusion:** The aim of telemedicine is to reduce disparities in healthcare access. The findings of this study show telemedicine has reduced racial disparities and provided greater accessibility for older groups. However, spatial and educational disparities are still noticeable. Research is necessary to examine how healthcare must address the socioeconomic heterogeneity in telemedicine by avoiding further disparities.

## Introduction

Novel Coronavirus Disease (COVID-19) is an infectious disease that has emerged worldwide by creating a pandemic in early 2020 (1). The COVID-19 Pandemic could be a significant health issue for the elderly population and individuals with pre-excising chronic conditions (e.g., hypertension, heart problems, and diabetes), who are more susceptible to developing the disease and have a higher risk of death (1, 2). In response to the pandemic, social distasting has played an important factor in slowing down the virus’s transmission. Telehealth is the delivery of health care services by health care professionals and currently is an essential tool in providing health care while providing patient and providers safe from COVID-19 (5, 6). Therefore, health care services have increased the use of telehealth medicine (3,4). The purpose of this study is to examine factors associated with access to telemedicine and scheduling before and after COVID-19 based on sociodemographic factors and the type of chronic disease. This study assesses access and scheduling telemedicine by age-group (e.g., 18-44,45-64, and 65+), race (e.g., White, Black, Hispanic, and other races), sex(males vs. females), educational status (high school or less, some college, bachelor’s degree or above), and urbanization (metropolitan vs. non-metropolitan). Research and Development Survey (RANDS) is longitudinal data that asks the participants in two points of time (June and August), which allows us to compare accessing and scheduling before and after COVID-19 and the progress of using telemedicine. This study, additionally, compares patients using telemedicine-based on chronic disease types to see which types of chronic diseases are more likely to be adopted for telemedicine faster in crisis moments.

Recent research has reported that during the COVID-19 pandemic, there are significantly lower levels of telehealth medicine usage among Black patients and the older population than their White counterparts (7). Instructional factors such as discrimination and segregation significantly affect the ability to access healthcare in an unfortunate event such as the COVID-19 pandemic. In addition, increase social vulnerability, including racial disparities, is associated with health inequalities (8). For example, one study has shown that COVID-19 exposure and death were higher among African American populations because they are disproportionately affected by multiple chronic diseases (9). A recent study in 2020 indicates that half of the sample were willing to use videoconferencing visits, and individuals who were older than 65 years (OR, 0.51) or had less education (OR, 0.37) were less likely to express willingness to have a telehealth appointment (10). Previous research shows that it is highly likely to have health disparities in non-metropolitan areas, and rural hospitals were less likely to use telehealth systems, such as checking patients’ health information online (11). Telehealth services are mostly for non-life-threatening health care; thus, there is a variation between the types of diseases in telemedicine. Dena (2020) shows individuals are more likely to have telehealth appointments for cardiovascular or metabolic disorders, mental health problems than autoimmune and respiratory diseases. Previous research examines the relationships between telemedicine and health disparities, including a few demographic measures, while this research includes these factors comprehensively in a longitudinal method.

In the following sections, the changes and progress of accessing and scheduling telemedicine in the first six months of the COVID-19 outbreak will be presented.

## Data and Results

This research uses data from the Research and Development Survey (RANDS) to administer survey question evaluation and statistical research. RANDS is a continuous range of studies for methodological research at the National Center for Health Statistics (NCHS). This research includes experimental evaluations of telemedicine access and uses for two rounds of RANDS during COVID-19. Data collection for the first wave occurred between June 9, 2020, and July 6, 2020, and data collection for the second wave was between August 3, 2020, and August 20, 2020. In the following sections, the percentage of telemedicine access before and after the pandemic and scheduling are presented.

**Table 1.** Summary of the Measures

|  | June | August |
| --- | --- | --- |
| **Age group** |  |  |
| 18-44 years | 38.41 | 37.03 |
| 45-64 years | 35.07 | 34.81 |
| 65 years and over | 26.51 | 28.14 |
| **Race** |  |  |
| White non-Hispanic | 66.4 | 68.2 |
| Black non-Hispanic | 11.95 | 11.53 |
| Other non-Hispanic | 7.79 | 7.71 |
| Hispanic | 13.85 | 12.54 |
| **Sex** |  |  |
| Male | 43.66 | 43.33 |
| Female | 56.34 | 56.67 |
| **Education** |  |  |
| High school graduate or less | 19.06 | 18.45 |
| Some college | 37.6 | 37.24 |
| Bachelor's degree or above | 43.34 | 44.3 |
| **Urbanization** |  |  |
| Metropolitan | 89.15 | 88.9 |
| Non-Metropolitan | 10.85 | 11.1 |
| **Provider offers telemedicine** |  |  |
| Yes | 36.6 | 37.1 |
| No | 46.9 | 48 |
| Do not know | 5.2 | 5.4 |
| No usual place of care | 11.3 | 9.5 |
| **Provider offered telemedicine prior to pandemic** |  |  |
| Yes | 14.1 | 14.1 |
| No | 61.7 | 62.6 |
| Do not know | 12.9 | 13.8 |
| No usual place of care | 11.3 | 9.5 |
| **Scheduled one or more telemedicine appointments** |  |  |
| Yes | 24.2 | 24.3 |
| No | 17.3 | 17.9 |
| No telemedicine available | 58.5 | 57.8 |

In June 2020, the first wave was asked (n= 6786), and the second wave was collected in August (n=5972). Three different questions were asked concerning telemedicine before and after COVID-19. The questions are 1) did the provider offer telemedicine before the pandemic? 2) does the provider offer telemedicine during the pandemic? And 3) have the participants schedule telemedicine appointments? The data includes age group, race, sex, education, urbanization status, and scheduling appointments by chronic diseases during the COVID-19 pandemic. In the first wave, around 37% of the sample had access to telemedicine, and 47% reported no access. There was no considerable change in the second round; however, 48% of the sample did not access telemedicine. In both waves, 62 % of the participants reported providers did not have telemedicine services prior to the COVID-19 pandemic. One out of four of the participants scheduled at least a telemedicine appointment in both rounds. There is no specific change in the distribution of demographic indicators in both waves. More than 25% of the sample were 65 years and over. Whites were the majority of the sample (66%), and 56% of the respondents were females. Less than 20% of the sample had high school or less, and close to 90% of the respondents from metropolitan areas.

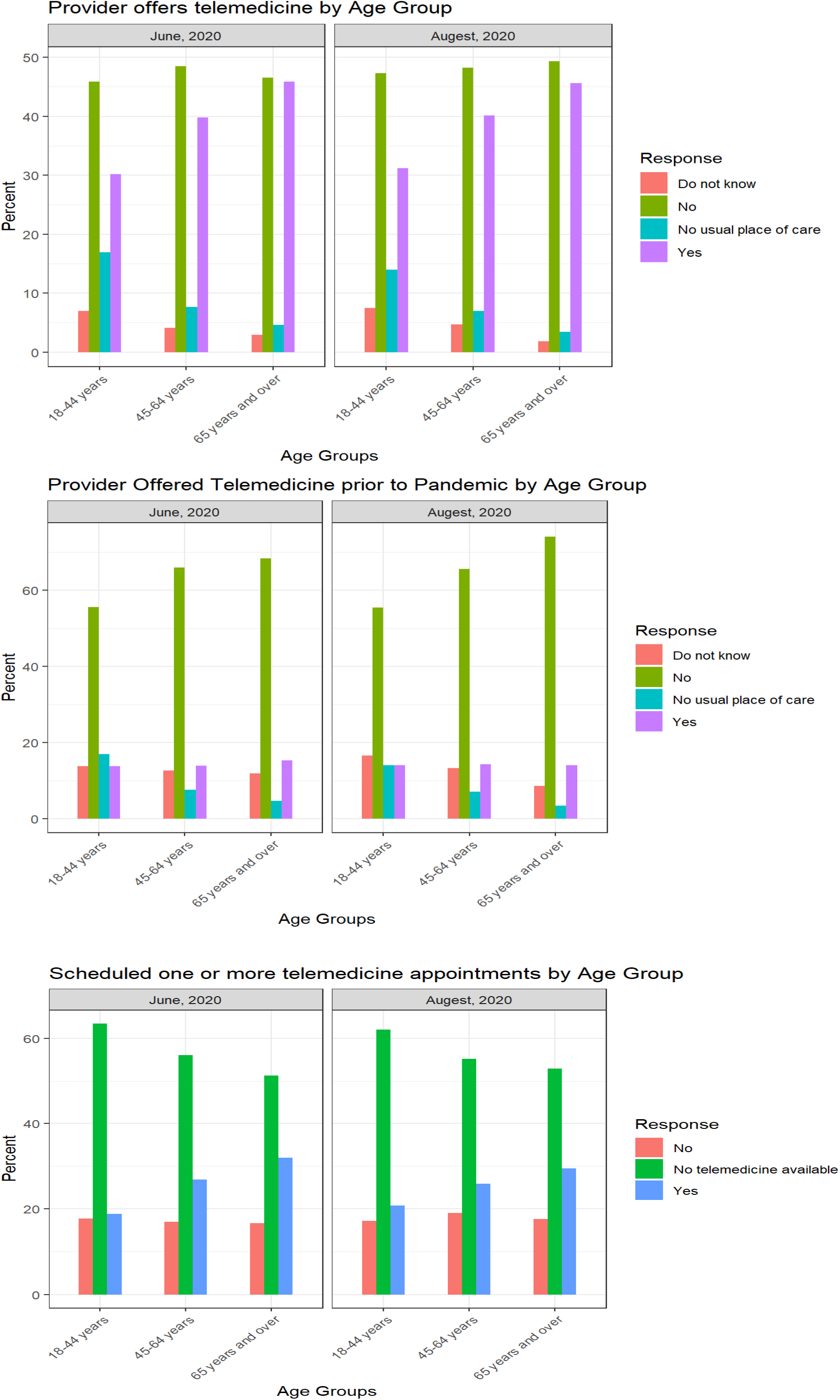

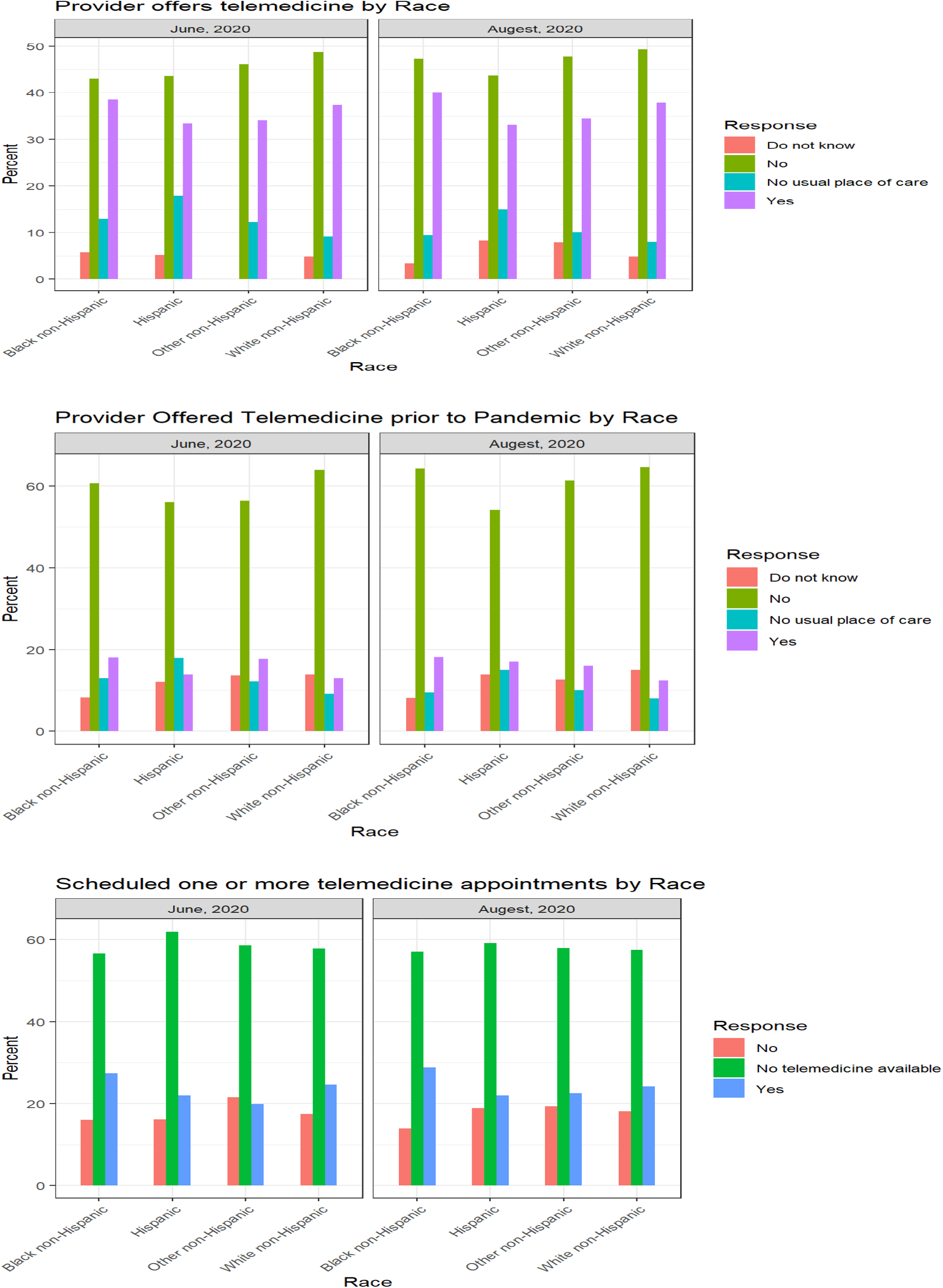

Results show that those older than 65 had higher access to telemedicine and had higher scheduling frequencies than other age groups. People who older than 65 reported 45% of providers offered telemedicine, while those between 18-44 reported 30% of providers had telemedicine services. Prior the pandemic, however, around 75 percent of people who were older than 65 reported the provider did not offer telemedicine. Blacks had the highest access to telemedicine services than other races(40%), and Whites had the next position(37%). Additionally, Blacks and Whites were more likely to visit doctors through telemedicine (close to 25%) than other races. Although there were no considerable differences between races in accessing telemedicine before the pandemic, Blacks had better outcomes than other races. Females were more likely to access and schedule during the COVID-19 pandemics than males, while females and males had the same performance before the pandemic. The results indicate 40 % of those with academic education reported accessing telemedicine, which was the highest percentage among other educational levels. Also, people with lower educational levels had higher rates of accessing telemedicine in August than June. Before the pandemic, there were no significant differences in accessing telemedicine based on educational level.

Although urbanization did not change people’s access to telemedicine, people who lived in non-metropolitan had slightly lower access in August than June. In the first round, participants in non-metropolitan areas reported 36% of providers offer telemedicine, while the access decreased 6% in August. This reduction was the same for scheduling for non-metropolitan regions in August. The findings indicate there was a variation in chronic disease types offered by providers during COVID-19. More than 55% of people diagnosed with diabetes could use telemedicine, while providers offer services around 40% for other chronic diseases. By comparison two rounds, the results show no change in the second round in providing telehealth by health care. More than 40 % of respondents who had diabetes reported at least one appointment through telemedicine, while 30% of patients with other chronic diseases had an online session. There was not a difference in offering telemedicine sessions for patients with the chronic disease before the pandemic.

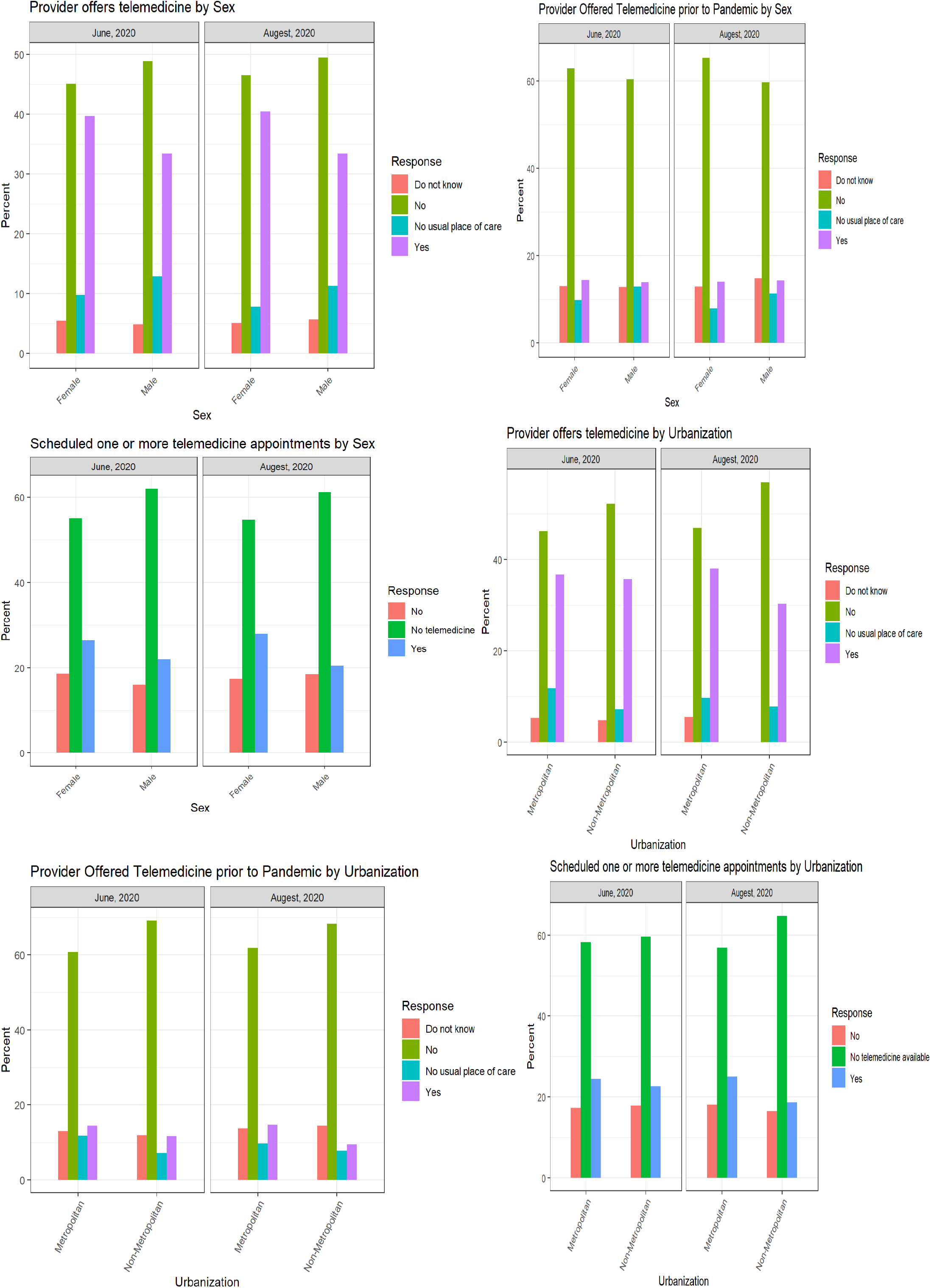

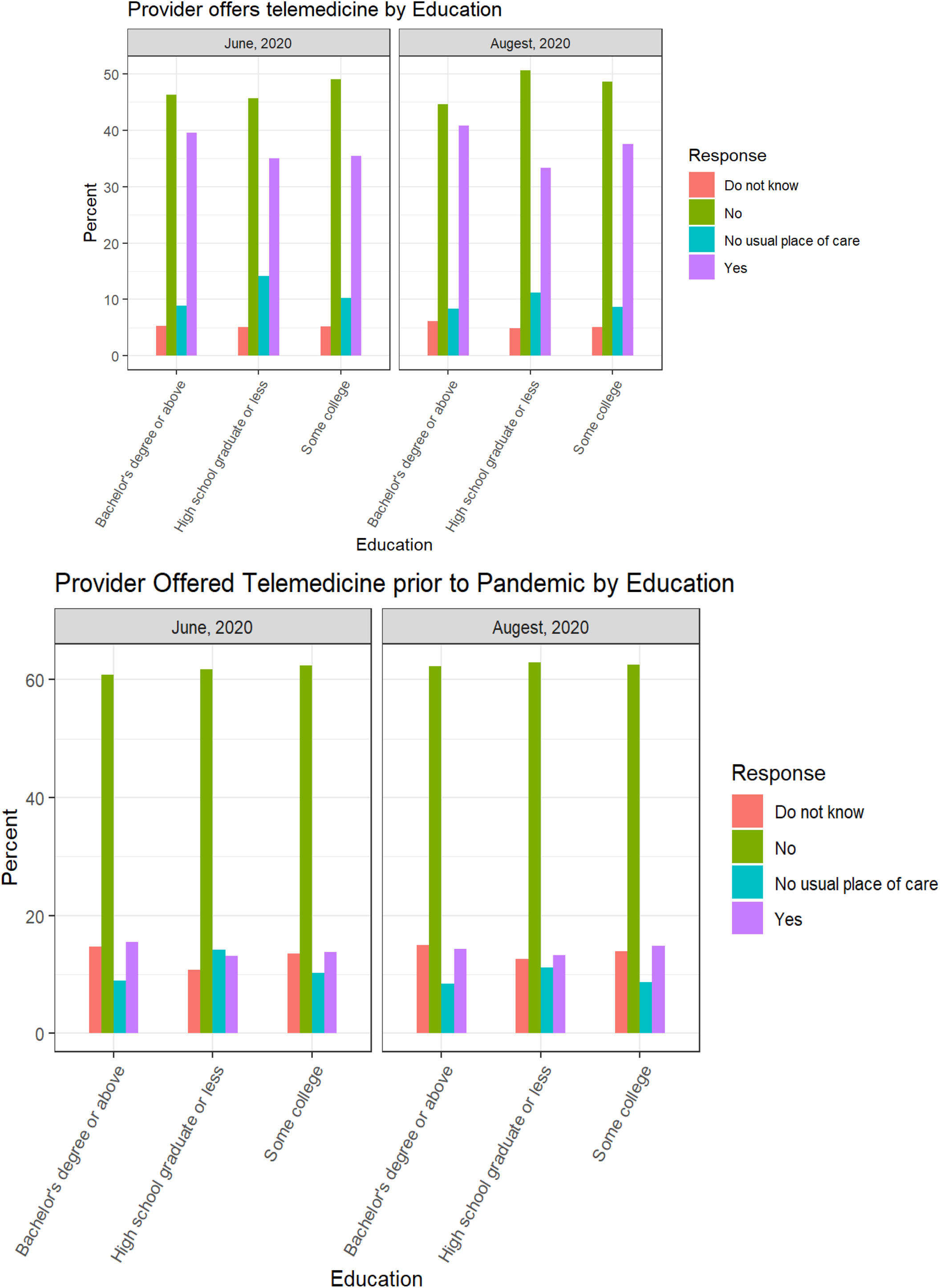

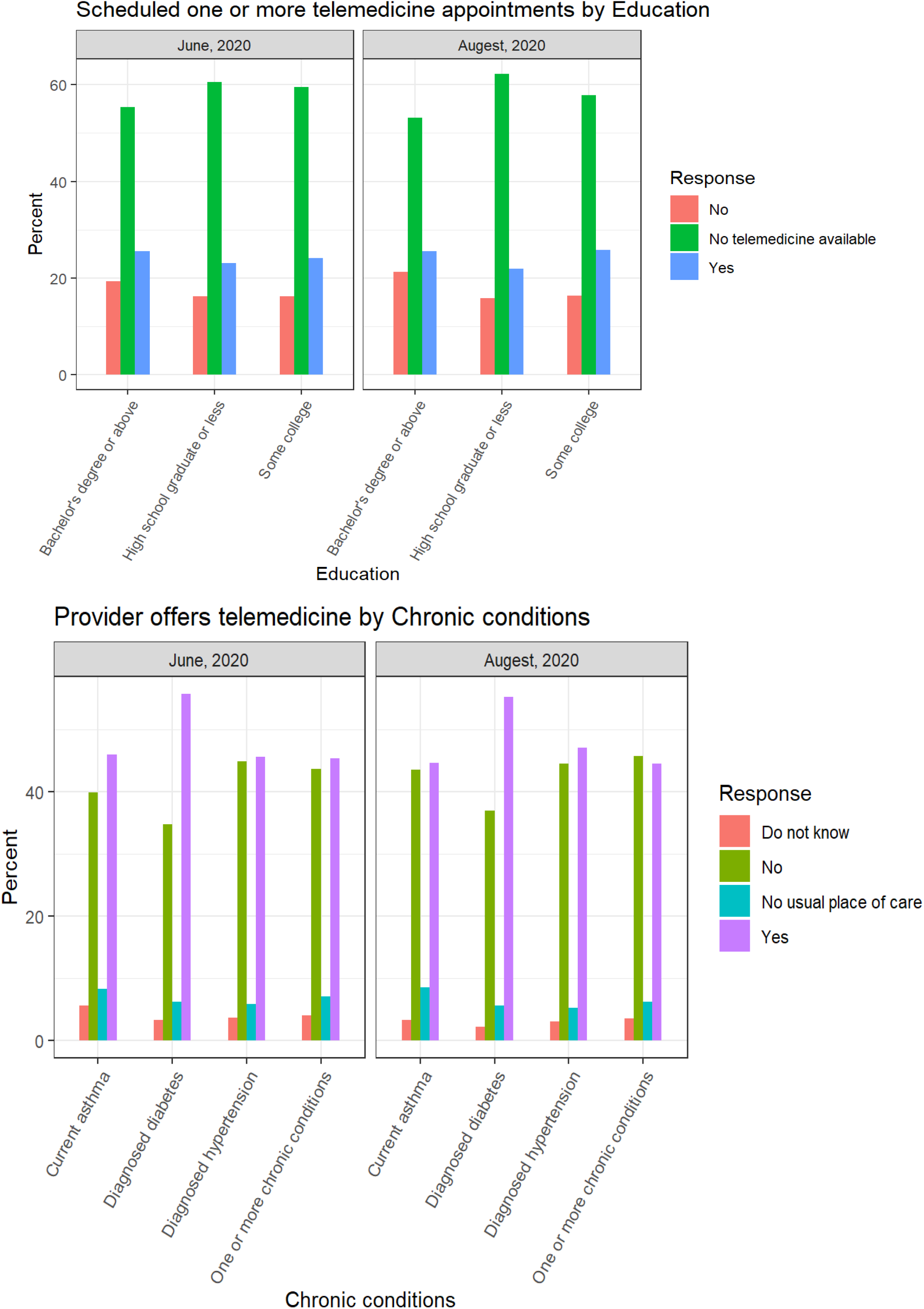

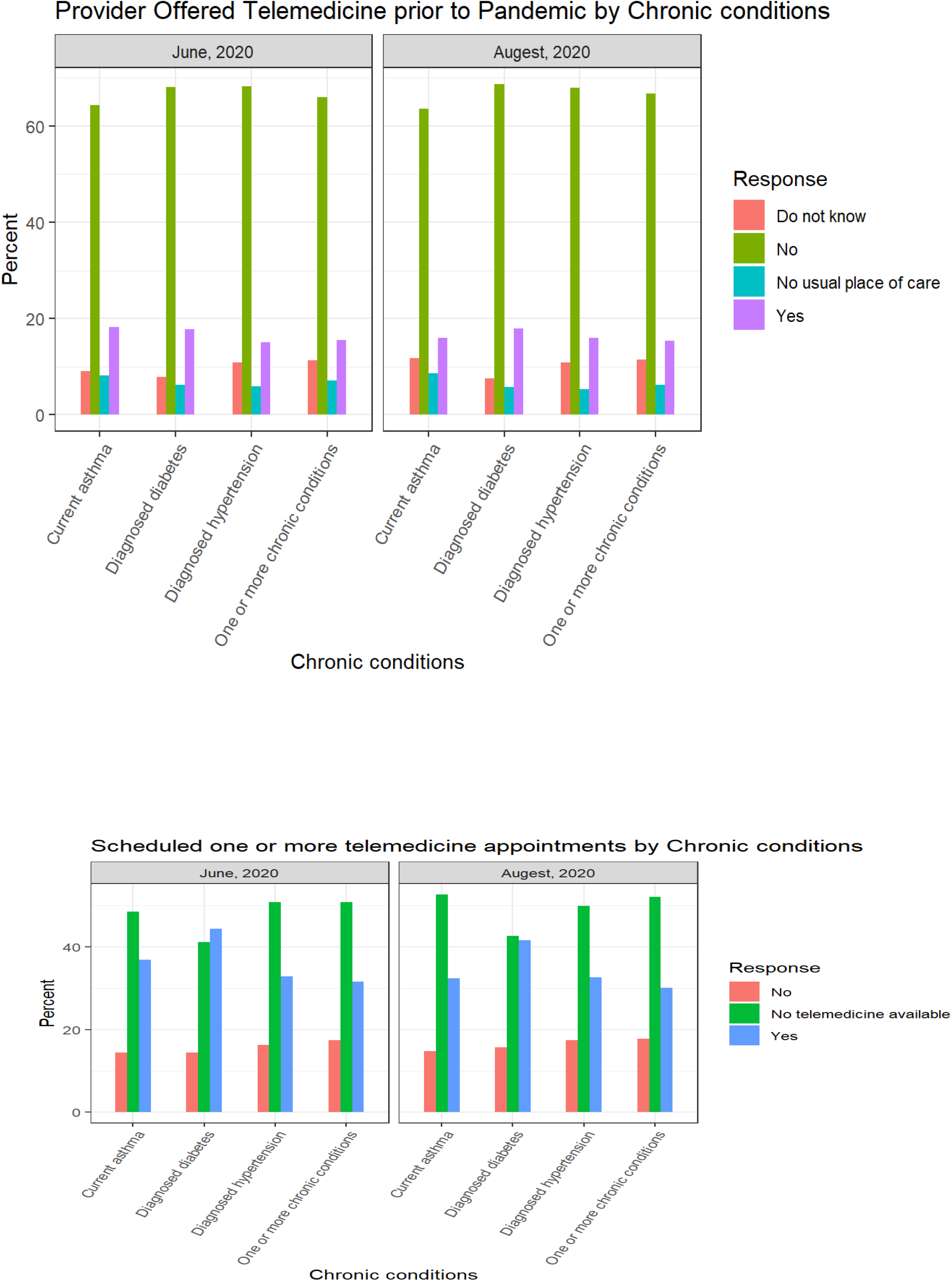

In sum, although older people had the lowest accessibility of telemedicine before the pandemic, they have the highest access and scheduling telemedicine during the pandemic (ref). Blacks reported better access to telemedicine than other races. Males and females had the same access to telemedicine before COVID-19; however, females had higher access and scheduling after COVID-19. Before the pandemic, education was not a source of disparity to access to telemedicine, while people with higher have more access and scheduling to telemedicine than those with lower educational levels after the pandemic.

Regarding urbanization and telemedicine, it was expected to have higher access to telemedicine in the second wave, however, non-metropolitan residents reported lower access than the first wave. This study shows providers offered higher services for diabetes than other chronic. The mechanisms of this study cannot explain why patients with diabetes had more scheduling and access to telemedicine, however; it is possible that some kinds of disease are more effective to follow up by telemedicine than other chronic diseases.

## Discussion

In a longitudinal survey of US residents, this study compares accessibility and scheduling telemedicine in two points of time, including before COVID-19, June 2020, and August 2020. we found a 22% increase in offering telemedicine in six first month of the COVID-19 pandemic. 36% of the respondents reported access to telemedicine, and close to 25% of them had at least one telemedicine appointment. Access to remote healthcare services is a promising solution for providing health care during the COVID-19 pandemic for those unable to visit providers in person, which is essential for stay-at-home orders. Although the period of data collection by RAND for the second wave was short (near two months), it would be promising providers increased their remote health care services during such a critical moment like the COVID-19 pandemic. However, our finding shows almost no change in providing telemedicine between June and August. The data indicates just a 0.5% and 0.1% increase in accessing telemedicine, and scheduling in August than June, respectively.

A primary aim of telemedicine is to reduce disparities in health care access, and the current research examines the access and scheduling of this digital tool based on demographic and socioeconomic indicators. The study by Dena (2020) examines the use of telehealth during the initial month of COVID-19, which shows those aged 18–44 years are more likely to have telehealth than older age groups, and the percentage of total visits was greater among younger than older adults. However, the current study’s findings indicate that the older group had a lower percentage of access before COVID-19, while they had more accessibility and scheduling telemedicine than other age groups. This result is surprising, as older age groups tend to be less engaged in digital technology, and telemedicine has not been a substantial barrier for older people anymore. Therefore, older people are more likely to adapt themselves to using remote health care services, and they have been more comfortable with technology and telehealth than in the past.

Another important finding in this study shows a reduction of disparities among Blacks and Whites by using telemedicine. Our study indicates that Blacks are more likely to use and access telemedicine during the COVID-19 pandemic than other races, which can show how telemedicine can be a vital opportunity to reduce disparities. Previous study discuss that Blacks were more likely to report they did not know how to use videoconferencing visits (10). Although the mechanism of this study cannot confirm this contradiction, this difference can be due to the type of telehealth or diseases. Future studies should concentrate on racial disparities on different types of telemedicine and diseases. In consistent with previous research, our results show that higher education and living in metropolitan areas are associated with higher using of telemedicine during COVID-19. Disparities in using telehealth are crucial to distinguish because telemedicine advocates frequently support telehealth’s ability to enhance access and decrease disparities in health services utilization. However, telehealth can be a source of disparities when people who would most benefit from it are the least likely to access and use it.

Despite evidence of higher access during the COVID-19 pandemic, telemedicine has yet remained at the periphery of the US healthcare system. Although the study compares different types of chronic diseases, and people who had diabetes were more likely to schedule or access telemedicine, the data was limited to chronic diseases. More information in other health sectors can be helpful to have more accurate pictures regarding telemedicine in the US. The second wave was asked in August, which was one of the first months of the COVID-19 onset. A longer duration of information also provides more accurate results. Lastly, we could not have interactions between socioeconomic and demographic measures to examine how racial disparities would differ in educational levels, sex, and age groups.

## Conclusion

Since the beginning of the COVID-19 pandemic health care system has changed its way of delivering care by providing telemedicine. This study examined factors associated with telemedicine access based on sociodemographic factors and the type of chronic disease associated with patients before and after the COVID-19 pandemic. Although telemedicine access has increased, there was no change in scheduling and access to telemedicine between June and August. Blacks and older ages had the highest access than other groups, while people with lower education or non-metropolitan areas had low access. Further research is necessary to examine how healthcare must address telemedicine’s socioeconomic heterogeneity by avoiding further disparities.

## Data Availability

The data is publicly available: https://www.cdc.gov/rdc/b1datatype/rdcrands.htm

THIS DATA IS AVAILABLE AT: https://www.cdc.gov/nchs/covid19/rands/telemedicine.htm#limitations

## Code Availability

The code to perform all analyses described in Methods and Usage Notes sections was written in R-3.6.2 and is publicly available at https://www.cdc.gov/rdc/b1datatype/rdcrands.htm

## Author Approval

All authors have seen and approved the manuscript.

## Author Contributions

Ali Roghani designed the study and implemented methods and analyses. Samin Panahi contributed to evaluate and develop the methods, and structure. All authors were involved in developing the ideas and drafting the paper.

